# Grouping theory of epidemiology

**DOI:** 10.1101/2021.06.11.21258661

**Authors:** Chaohui Zhang, Wenyu Ling

**Affiliations:** (Guangzhou, China)

**Keywords:** trend, grouping, differentiated treatment, influence value, contribution value, utility coefficient, control coefficient, vaccine, herd immunity

## Abstract

**Background (traditional theory):** there are general and principle models and calculation formulas for epidemics, mainly the basic reproduction number R_0_ of a certain epidemic disease such as COVID-19, the newly confirmed number C_n_=R_n-1_C_n-1_, The threshold of herd immunity is (R_0_-1)/R_0_.

**Innovation theory:** Based on the fact that there are groups of different tends (that is, different R_0_ or R_k_), a grouping model of epidemiology is proposed, and a complete and detailed calculation formula is given. The basic relationship is that the overall infection numbers of is equal to the sum of the infection number of groups, namely: R_t_=∑(f_k_R_k_), where f_k_ is the population proportion of each group, and R_k_ is the basic reproduction number of each groups. Its important application is the grouping strategy in which prevention and control measures are inclined to high-risk groups. The basic relationship is that the basic reproduction number of each group is equal to the product of the control coefficient of its measures and the original basic reproduction number, namely: R_k_=ζ_k_R_0k_. Compared with the traditional strategy of equal treatment, the grouping strategy has the characteristics of high efficiency, low consumption, and low threshold of herd immunity.

## Introduction

Since the outbreak of the covid-19 epidemic for more than a year, apart from the success of vaccine research and development and the success of a few countries, the global response has been in chaos. The important reason lies in the backwardness of epidemic analysis and prevention and control measures and the lack of theory, which makes the response lack basis, there are serious differences and controversies. And even the so-called herd immunity strategy of natural infection can be popular and cause great harm. Therefore, filling the gaps and proposing effective theories is the top priority. This article discusses this. In the “Principle Model of Epidemic Infection Based on Differentiating R and Differential Treatment Strategies” in November 2020, I have proved that there are different trend groups in the population, and proposed a simple group analysis model and group prevention and control strategy. This article further demonstrates the principles, formulas and strategies of the model, which are referred to as grouping model and grouping strategy, collectively referred to as grouping theory.

## Grouping models and formulas

### Parameters and representations

For simplicity, the basic reproduction number is represented by R, the original basic reproduction number is R_0_, and the basic reproduction number of the k group is R_k_. The subscript of the symbol parameter can include the letter k and the number n. K represents the potential group, namely k=a, b, c…, where k=t represents the whole, and n represents the generation, such as C_an_、C_b(n-1)_ 、C_tn_、A_tn_、A_t15_. When there is no need to distinguish between generations, subscripts do not use n, such as R_a_、R_b_、R_t_、f_c_、f_d_、r_t_、r_a_. When there is no grouping, the subscript does not use k, such as C_n-1_ 、C_0_、A_n_。f_k_ represents the population proportion of the group, so f_a_+f_b_+f_c_+……=f_t_=1.

There are many factors that affect the R, mainly innate immunity, physical fitness and occupation (the degree of contact with the population, especially the infected carriers). The first two are closely related to age, and retirement is related to age. As a principle analysis, this article is divided into groups by age.

### The basic relationship of the principle model (without grouping)

When without grouping, subscript k is no need, and its formula is the traditional principle formula:

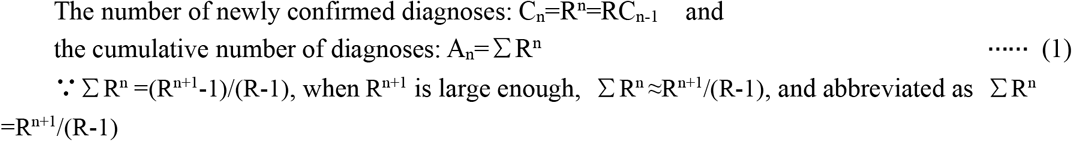

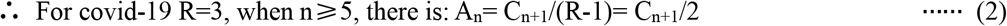

### Grouping principle model and formula

Based on the existence of different R, the grouping principle model introduces the basic reproduction number R_k_ of each group. The following takes 3 groups a, b, and c as an example. The R of each group is R_a_, R_b_, R_c_, and the basic reproduction numbers of the three groups are: the strong group R_a_, the medium group R_b_, the weak group R_c_, and the whole is R_t_, C_tn_ is added as a whole. The principle is that when each group is not independent, the source of infection C_k(n-1)_ is allocated according to the population proportion f_k_ of each group, that is: C_k(n-1)_= f_k_C_t(n-1)_, and then each group infect according to its own R. Therefore, the formula and its derivation are:

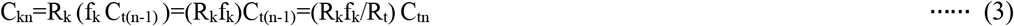

Among them, I_k_=R_k_f_k_=R_t_C_kn_/C_tn_ is called the impact value of group k on the epidemic, namely R. Note: ∑R_k_=R_a_+R_b_+R_c_≠R_t_=3

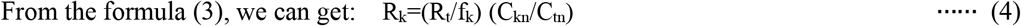

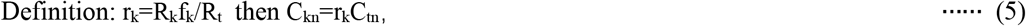

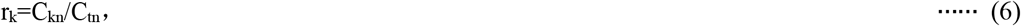

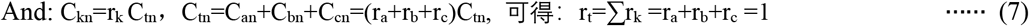

It can be seen that r_k_ is the infection distribution coefficient of the group, and there are:

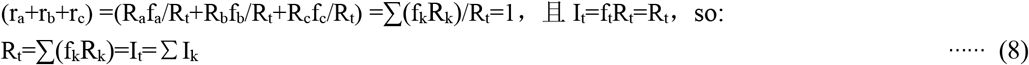

### Practical calculation formula based on statistical data

The above formula uses C but not A, such as (4) formula R_k_=(R_t_/f_k_) (C_kn_/C_tn_), but the data of C is very unstable in statistics, and the error is large, while A is relatively stable and the error is small, so when practical The formula for A is often used to replace the formula for C. The specific derivation is as follows:

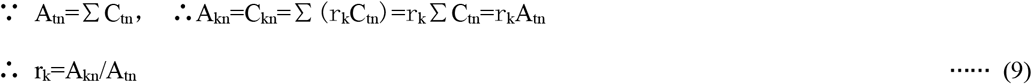

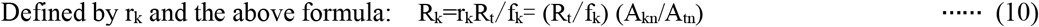

From the above, it can be obtained (subscript n omitted):

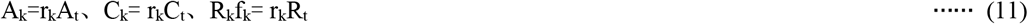

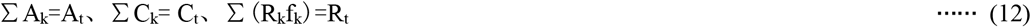

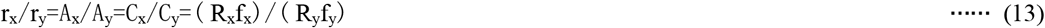

Note that each group is not a closed body infected by independent sources of infection C_k0_ (or C_k(n-1)_) and R_k_. If it is regarded as a closed body, the principles, formulas, and results will be completely different. For example, the trend of each group will not be so different, otherwise the weak group with R=7 will be all infected early, and the strong group with R less than 1 will not need to prevent and control because it will not be epidemic.

## Principles, formulas and analysis tables of grouping strategies

### Control measures and their effects and parameters

The assumptions and data in Table 1 and the following analysis are inaccurate, but because this article is the principle, trend and principle formula, especially basic, the main formula does not include specific data such as C_n_、C_n-1_、A_k_、A_kn_, etc., so Not affected by them. However, specific and accurate data, parameters and analysis are required in practical applications, such as R, A, r, and ζ. Therefore, the following tables and analyses use simplified data.

**Table 1.** Covid-19 data sheet for four age groups

| Gr | Age | $f_k$ | $R_k=R_0 r_k/f_k$ | $I_k= f_k R_k$ | $r_k=f_k R_k/R_t$ |
| --- | --- | --- | --- | --- | --- |
| a | 0 ~ 29 | 0.28 | 0.6 | 0.17 | 0.06 |
| b | 30 ~ 49 | 0.27 | 2 | 0.54 | 0.18 |
| c | 50 ~ 69 | 0.28 | 4 | 1.1 | 0.37 |
| d | $\geq 70$ | 0.17 | 7 | 1.2 | 0.39 |
| t | Whole | $f_t=\sum f_k=1$ | $R_t=R_0=3$ | $f_t R_t=R_t=\sum (f_k R_k)$ | $r_t=\sum r_k=1$ |

Wherein: Gr: Group; $f_k$ : Proportion of population Overall; $I_k$ : Impact value; $r_k$ : infection distribution coefficient .

Generally, management and control measures are classified into different levels. Here, level 1 and level 2 are used to indicate gradual escalation of measures. The parameters changed after the measures are taken are still represented by the original names and symbols, while the parameters before the measures are called the original parameters and are indicated by subscript 0. For example, the original basic reproduction number of the covid-19 is R_0_=3, and the basic infection after the measures The number is R_t_. Define ξ as the utility coefficient of the control measures, and the reduction of the basic reproduction number R is ΔR, then ΔR=ξR, ΔR_0_=ξR_0_,ΔR_k_=ξR_k_. Define ζ as the control coefficient, then R_t_=ζR_0_, because R_t_=R_0_-ΔR_0_=(1-ξ)R_0_=ζR_0_, so ζ=(1-ξ), and the values of the two coefficients are both between 0 and 1. Accordingly, there are:

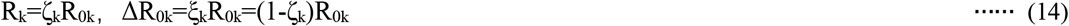

The larger the ξ, the smaller the ζ. For example, level 0 is uncontrolled natural infection, ξ=0, ζ=1. level 1 is conventional prevention and control measures, such as wearing masks, washing hands frequently, etc., tracking and monitoring, isolation of carriers, ξ=0.7, Z=0.35. Level 1-is Level 1 measures for poor execution. For example, it is only recommended, tracking and isolation are also poor, ζ=0.4∼0.6. Level 2 is Level 1 plus distance, ζ=0.3. Level 2-, ζ=0.35∼0.45. Level 3 is level 2 plus some special facility restrictions or even shut down, ζ=0.25. Level 3-, ζ=0.3∼0.4. Level 3+ means home isolation, ζ=0.25∼0.15. Level 4 is level 3 plus general facilities such as schools, factories, shops, ζ=0.15. level 4-, ζ=0.2∼0.3. level 5 is level 4 plus home isolation, severely restricted daily life, economic activities severe stagnation, ζ=0.1. Level 5+ means the area is blocked, ζ=0.05. Let ζ_k_ denote ζ of the k group. Customs blockade is an overall special measure. When ξ=(R_0_-1)/R_0_, R_t_=1, and ζ=1/R_0_ at the same time, the curve of newly added diagnoses is flat. If the curve flattening is achieved by a sufficient proportion of the immunized population, it is called herd immunity.

### Production and analysis of management and control analysis table

The original parameters R_0_ and f_k_ are calculated from the statistical data, and then r_k_=A_kn_/A_tn_ is obtained from A_tn_ and A_kn_, and then R_k_=R_t_r_k_/f_k_, thus the original parameter table 1 is established. In addition, it is necessary to study statistics to obtain the control coefficient ζ of various control measures. They are the basis for the analysis, based on which the management and control analysis tables such as Tables 2, 3, 4, etc. can be established, and the herd immunity analysis table for vaccines can also be established 。

**Table 2.** Analysis table under the same control measures. Let R_t_=1, then ζ_t_=0.33. Since ζ_k_ is the same, ζ_k_=ζ_t_=0.33.

| Gr | Age | $f_k$ | $R_{0k}$ | MNS | $\zeta_k$ | $R_k$ | $I_k$ | $r_k$ |
| --- | --- | --- | --- | --- | --- | --- | --- | --- |
| a | 0 ~ 29 | 0.28 | 0.6 | L3- | 0.33 | 0.19 | 0.06 | 0.06 |
| b | 30 ~ 49 | 0.27 | 2 | L3- | 0.33 | 0.66 | 0.18 | 0.18 |
| c | 50 ~ 9 | 0.28 | 4 | L3- | 0.33 | 1.3 | 0.37 | 0.37 |
| d | $\geq 70$ | 0.17 | 7 | L3- | 0.33 | 2.3 | 0.39 | 0.39 |
| t | Whole | $f_t=1$ | 3 | Closure | 0.33 | 1 | 1 | $r_t=1$ |

Wherein: MNS: control measures; L3-:Level 3-; $R_k=\zeta_k R_{0k}$ ; $I_k=f_k R_k$ ; $I_t=f_t R_t=R_t=\sum(f_k R_k)$ ; $r_k=f_k R_k/R_t$ ; $r_t=\sum r_k=1$

**Table 3.** Analysis table of R_t_=1 under different control measures

| Gr | Age | $f_k$ | $R_{0k}$ | MNS | $\zeta_k$ | $R_k=\zeta_k R_{0k}$ | $I_k=f_k R_k$ | $r_k=f_k R_k/R_t$ |
| --- | --- | --- | --- | --- | --- | --- | --- | --- |
| a | 0 ~ 29 | 0.28 | 0.6 | L1- | 0.5 | 0.3 | 0.08 | 0.08 |
| b | 30 ~ 49 | 0.27 | 2 | L2- | 0.4 | 0.8 | 0.22 | 0.22 |
| c | 50 ~ 9 | 0.28 | 4 | L3- | 0.3 | 1.2 | 0.34 | 0.34 |
| d | $\geq 70$ | 0.17 | 7 | L3- | 0.3 | 2.1 | 0.36 | 0.36 |
| t | Whole | $f_t=1$ | 3 | Closure | $\zeta_t=R_t/R_{0t}=0.33$ | $R_t=1$ | $R_t=1$ | $r_t=\sum r_k=1$ |

**Table 4.** Analysis table under the Differential measures, R_t_=0.8

| Gr | Age | $f_k$ | $R_{0k}$ | MNS | $\zeta_k$ | $R_k=\zeta_k R_{0k}$ | $I_k=f_k R_k$ | $r_k$ | $\xi_k$ | $f_k R_{0k}$ | $Q_k$ |
| --- | --- | --- | --- | --- | --- | --- | --- | --- | --- | --- | --- |
| a | 0 ~ 29 | 0.28 | 0.6 | L2 | 0.3 | 0.2 | 0.06 | 0.08 | 0.7 | 0.17 | 0.12 |
| b | 30 ~ 49 | 0.27 | 2 | L2 | 0.3 | 0.6 | 0.16 | 0.22 | 0.7 | 0.54 | 0.38 |
| c | 50 ~ 9 | 0.28 | 4 | L2 | 0.3 | 1.2 | 0.34 | 0.43 | 0.7 | 1.12 | 0.78 |
| d | $\geq 70$ | 0.17 | 7 | L3+ | 0.2 | 1.4 | 0.24 | 0.30 | 0.8 | 1.19 | 0.95 |
| t | Whole | $f_t=1$ | 3 | Closure | $\zeta_t=0.27$ | $R_t=0.8$ | $R_t=0.8$ | $r_t=1$ | 0.73 | 3 | $Q_t=2.2$ |

Among them: $I_{0k}=f_k R_{0k}$ , $Q_k=\xi_k I_{0k}$ , $\zeta_t=R_t/R_{0t}$ , $Q_t=\sum Q_k=R_{0t}-R_t$

**Table 5.**
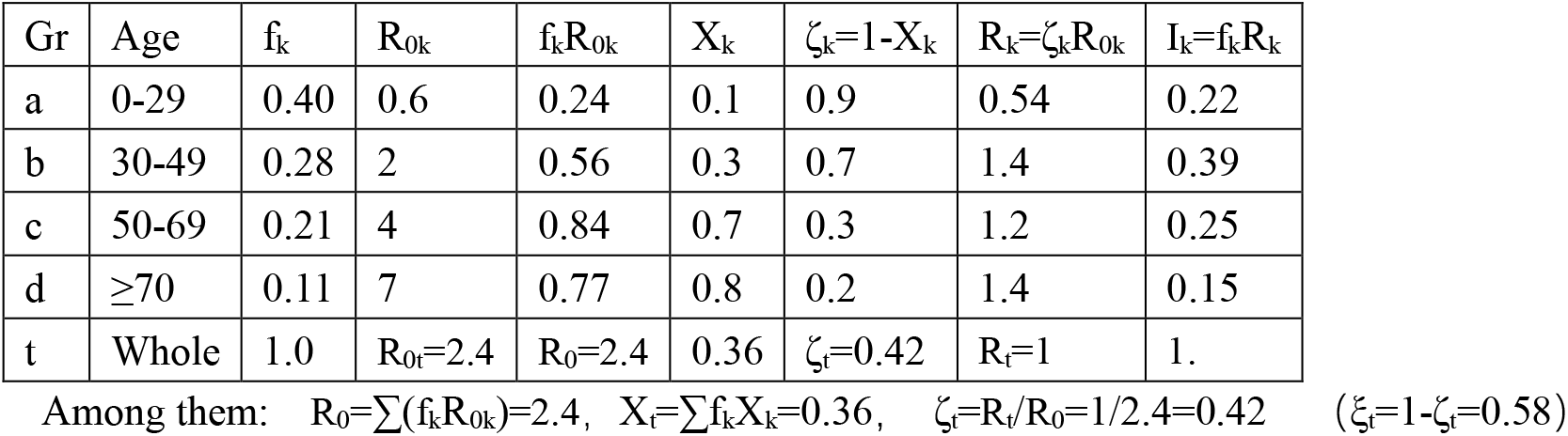
Herd immunity analysis of vaccination under the grouping strategy. Among them: ζ_k_=1-ξ_k_=1-X_k_

| Gr | Age | $f_k$ | $R_{0k}$ | $f_k R_{0k}$ | $X_k$ | $\zeta_k=1-X_k$ | $R_k=\zeta_k R_{0k}$ | $I_k=f_k R_k$ |
| --- | --- | --- | --- | --- | --- | --- | --- | --- |
| a | 0-29 | 0.40 | 0.6 | 0.24 | 0.1 | 0.9 | 0.54 | 0.22 |
| b | 30-49 | 0.28 | 2 | 0.56 | 0.3 | 0.7 | 1.4 | 0.39 |
| c | 50-69 | 0.21 | 4 | 0.84 | 0.7 | 0.3 | 1.2 | 0.25 |
| d | $\geq 70$ | 0.11 | 7 | 0.77 | 0.8 | 0.2 | 1.4 | 0.15 |
| t | Whole | 1.0 | $R_{0t}=2.4$ | $R_0=2.4$ | 0.36 | $\zeta_t=0.42$ | $R_t=1$ | 1. |

Among them: $R_0=\sum(f_k R_{0k})=2.4$ , $X_t=\sum f_k X_k=0.36$ , $\zeta_t=R_t/R_0=1/2.4=0.42$ ( $\xi_t=1-\zeta_t=0.58$ )

### Original data sheet

Table 1 is based on the Italian epidemic data on March 11, 2020, but simplified it to four age groups, removes the death and A_kn_ items, and increases the infection distribution coefficient of each group r_k_=A_kn_/A_tn_, R_k_=R_t_r_k_/f_k_, and impact value I_k_= f_k_R_k_. Although the data is not statistically complete and accurate, it has the significance of an example in principle. Therefore, the analysis and demonstration in principle can be carried out with this data.

### The impact value of the group on the epidemic and its key

The number of newly confirmed diagnose C_k_=R_k_f_k_C_t_ in different groups is different, so the impact on the overall number of infections is different. Since C_t_ is the same for each group, the impact can be described intuitively by the group’s impact value I_k_=f_k_R_k_ or the proportional distribution coefficient r_k_=f_k_R_k_/R_t_. There are also related relations: I_t_=R_t_ =∑I_k_=∑(f_k_R_k_), ∑r_k_=r_t_=1 (Because the impact value I_k_ varies with R_t_, and the sum of r_k_ is a constant 1, sometimes r_k_ is more convenient to compare the influence.). The influence assessment is mainly to compare I_k_, but the key is R_k_. For example, in Table 1: the f_k_ of group a and group c are the same, but R_k_ is very different, the ratio of the two I_k_ is the ratio of R_k_: R_a_/R_c_=0.17/1.1=0.6/4=0.15; group a and group d The population ratio is 1.6, but the ratio of R_k_ is 0.09, so the ratio of I_k_ is 0.28*0.6/(0.17*7)=0.14, and the influence is much smaller. The f_k_ of the c group is much larger than d group, but the R_k_ is much smaller, so the impact value is similar. To sum up, the influence on the epidemic mainly depends on R_k_, followed by f_k_. Since f_k_ is fixed and R_k_ changes with measures, control measures should be tilted towards groups with large R_k_.

### Contribution value of the group to the measure

The reduction of the overall basic reproduction number after taking measures is ΔR= R_0_-R_t_. The contribution of each group to ΔR is different. Define Q_k_ as the contribution value of the group to ΔR. Its definition and relational expression are (Note: the proof and derivation are omitted here. Note: R≠∑R_k_, ΔR≠∑ΔR_k_):

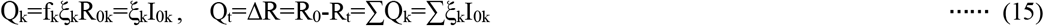

It can be seen that the contribution value of the measure is proportional to ξ_k_, 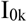 or ξ_k_, f_k_, 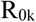. Since f_k_ is a constant, the key is ξ_k_ and 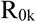. Therefore, if you want the strategy to achieve greater benefits, you should take measures with greater ξ_k_ for groups with greater impact value 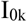, especially greater 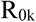, at this time the measure will have a multiplied contribution value. For example, in Table 4, group d only accounts for 17% of the population, but its 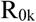 is as high as 7, and the utility coefficient is 0.8. So although its utility coefficient ξ_d_ is only 1/7 higher than the 0.7 of other groups, its contribution value of 0.95 is 20% higher than the 0.78 of the c group. c group has a population of 1/3 more than him, and 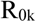 is Up to 4. It is even 8 times higher than group a, whose population is 1/3 larger than his.

## Grouping strategy based on grouping model

Based on the fact that different groups have different trends, namely R, the prevention and control of the epidemic should adopt a differentiated grouping strategy. The following examples discuss its principles.

### non-differentiated control strategy and its analysis

Still taking the situation of the outbreak in Italy at that time and R=3 as an example, the top priority at this time is to control the spread and reduce R to less than 1. Therefore, it is necessary to adopt level 3-, and add customs blockade measures. According to this measure, Table 2 is obtained, where 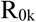 is the original R_k_ of Table 1, and R_k_ is the basic reproduction number under control. It has two more than Table 1: control measures and ζ_k_. Because the measures are the same, the ζ_k_ of each group is the same, and they are all equal to ζ_t_, so r_k_ does not change, but ζ_t_ changes with R_k_.

This is similar to a measure taken by most countries, especially Western countries, when the epidemic is severe. Although the control measures are quite stringent and the epidemic is controlled, people over 70 are still in a severe situation with a high infection rate R_k_=2.3, and high mortality rate. And the 0-29 year-old population with a low R_k_=0.6 is suffered from inefficient strict control, and so is subject to unnecessary strong negative effects.

### Differential treatment strategy and its analysis

If we treat them differently, and adopt incremental control measures for each group in the order of high and low trend, very good results will be achieved. At this time, the control measures can be set first, R_k_ and I_k_ can be introduced by ζ_k_, and r_k_ can be used as verification, and then the control measures can be adjusted to obtain the target R_t_. The following table shows the analysis of the differential treatment control measures, that have the same effect as the non-differential treatment control measures table 2, namely R_t_=1.

Specifically: the measures of strong groups a and b are level 1 and 2. but due to the poor execution of masks and distancing, the control coefficient ζ is only 0.5 and 0.4. The weaker groups c and d adopt stricter 3-,plus home isolation, and their control coefficient ζ reaches 0.3. So the result is R_t_=1. Although the restrictions on groups c and d have been stricter, their protection has also been strengthened. At the same time, the restrictions on groups a and b are greatly reduced, so the overall impact on the economy and life is much smaller.

If the enforcement for measures are differently, the control coefficients are different. For example, if the measures to wear mask are implemented well, then ζ=0.4, plus social distancing is maintained well, then ζ=0.3. For level 3+ (plus home isolation) with strong execution, ζ=0.2. At this time, R_t_=0.8, which is a very good downward curve. The analysis is shown in the following table. It adds several items such as contribution value. The last four items are used for the analysis of the principle, and may not be listed in practice.

## Analysis of Grouping Theory Applied to Vaccination

There is no doubt that the grouping theory can be applied to the herd immunity analysis and strategy of vaccination. Its basic principle is: the product of the inoculation rate X_k_ and the effective rate is the utility coefficient of vaccine prevention and control measures. What is interesting is that, based on intuition, countries generally implement a differentiated vaccination strategy, so which can be analyzed using grouping theory. The following table is based on a typical age distribution and vaccination rate distribution of the aging population in the United States. it makes a simplification: offset the invalid part of vaccines with the immunity rate formed by infection, so forming a 100% vaccine effective rate and zero infection immunity rate Simplified model, among them, the vaccination rate is the utility coefficient of vaccination. this table shows that according to its 95% vaccine effective rate, as long as the overall vaccination rate reaches 36%, herd immunity can be achieved. Considering simplification and other errors, it can be considered that if the vaccination has an overall coverage rate of 45%, herd immunity can be achieved. However, the traditional theory of non-grouping, only when the vaccine effective rate is 100%, and the vaccination rate reaches 67% the herd immunity can be achieved. If the effective rate is 90%, the vaccination rate must be 72%.

From March to May in the United States, Texas and Florida, which “should” have soared due to the adoption of full liberalization measures, but had the same decline in the epidemic as California and New York, which adopted the lockdown strategy. This puzzled experts.. According to guruin.com data, their (adults?) vaccination rates on May 25 were: 43% in Texas, 48% in Florida, 55% in California, and 54% in New York. According to traditional theory, this is far from reaching the 67% herd immunity requirement, and it is impossible to completely open up without increasing but falling. However, it can be easily explained by the analysis of the grouping theory as shown in the above table: their vaccination rates have reached or close to 36%, plus natural reserved epidemic prevention habits and measures, such as some people wear masks, customs sealing etc., so R is already less than 1. It is equivalent to achieving herd immunity.(Another important reason is that its relatively young and aging population structure makes its basic reproductive number R0=2.4 much smaller than 3, and the traditional herd immunity threshold is not 67% but 58%.)

## Concluding remarks

For Covid-19, there are significant and distinguishable groups. This fact determines that when the epidemic is severe, a different prevention and control strategy must be adopted. This is an important principle for prevention and control of the epidemic. Based on the above analysis, it can be seen that the grouping strategy not only doubles the result with half the effort, but also reduces or even avoids blockades and ensures economic operation. The non-differential treatment strategy is not only half the effort, but also huge losses. This article provides basic principles, formulas and analysis tables. The analysis table is just a principle model based on statistics and general experience. It includes a large number of simplifications and approximations, which are equivalent to ideal models and smooth curves of formulas. Therefore specific analysis and application are required in application. In actual prevention and control, the groups can be divided into more detailed groups, and different groups and strategies can also be used. For example, front-line medical staff are both high-risk groups and cannot be isolated, and they need high-intensity protection.

——End

## Data Availability

This manuscript does not cite such data.

## About the author

retired mechanical engineering teacher; Independent research, no tutors, no funds. **Email**

